# Tuberculosis biomarkers discovered using Diversity Outbred mice

**DOI:** 10.1101/2021.01.08.20249024

**Authors:** Deniz Koyuncu, M. Khalid Khan Niazi, Thomas Tavolara, Claudia Abeijon, Melanie Ginese, Yanghui Liao, Carolyn Mark, Adam C Gower, Daniel M Gatti, Igor Kramnik, Metin Gurcan, Bülent Yener, Gillian Beamer

**Affiliations:** Rensselaer Polytechnic Institute, Department of Electrical, Computer, and Systems Engineering, Troy, NY 12180; Wake Forest School of Medicine, Bowman Gray Center for Medical Education, Winston-Salem, NC, 27101; Tufts University, Cummings School of Veterinary Medicine, North Grafton, MA 01536; Kansas State University, College of Veterinary Medicine, Manhattan, KS 66506; Boston University Clinical and Translational Science Institute, Boston, MA 02118; The College of the Atlantic, Bar Harbor, ME 04609; Boston University, National Emerging Infectious Diseases Laboratories, Boston, MA 02118; Rensselaer Polytechnic Institute, Department of Computer Science, Troy, NY 12180

## Abstract

**Background:** Biomarker discovery for pulmonary tuberculosis (TB) may be accelerated by modeling human genotypic diversity and phenotypic responses to *Mycobacterium tuberculosis* (*Mtb*). To meet these objectives, we use the Diversity Outbred (DO) mouse population and apply novel classifiers to identify informative biomarkers from multidimensional data sets.

**Method:** To identify biomarkers, we infected DO mice with aerosolized *Mtb* confirmed a human-like spectrum of phenotypes, examined gene expression, and inflammatory and immune mediators in the lungs. We measured 11 proteins in 453 *Mtb-*infected and 29 non-infected mice. We have searched all combinations of six classification algorithms and 239 biomarker subsets and independently validated the selected classifiers. Finally, we selected two mouse lung biomarkers to test as candidate biomarkers of active TB, measuring their diagnostic performance in human sera acquired from the Foundation for Innovative New Diagnostics.

**Findings:** DO mice discovered two translationally relevant biomarkers, CXCL1 and MMP8 that accurately diagnosed active TB in humans with > 90% sensitivity and specificity compared to controls. We identified them through the two classifiers that accurately diagnosed supersusceptible DO mice with early-onset TB: Logistic Regression using MMP8 as a single biomarker, and Gradient Tree Boosting using a panel of 4 biomarkers (CXCL1, CXCL2, TNF, IL-10).

**Interpretation:** This work confirms that the DO population models human responses and can accelerate discovery of translationally relevant TB biomarkers.

**Funding:** Support was provided by NIH R21 AI115038; NIH R01 HL145411; NIH UL1-TR001430; and the American Lung Association Biomedical Research Grant RG-349504.

## Introduction

Tuberculosis (TB) remains a global health crisis. The disease is diagnosed in 8-10 million new patients and has caused 1-2 million deaths every year for many decades. This is nearly 5000 deaths per day, higher than the average daily death rate due to COVID-19 ^1^. Pulmonary TB accounts for 70-80% of TB cases, is contagious, and has a 40-70% case fatality rate if untreated ^2-5^. For decades, studying inbred laboratory mice provided critical insight into host responses to *Mycobacterium tuberculosis* (*Mtb)* that were controlled by single cell types and single genes. The field, however, is currently undergoing two shifts: 1) Inclusion of genetically heterogenous animal models to identify factors that control *Mtb-*induced lung damaging inflammation in immune competent hosts; and 2) Use of new methods to identify biomarkers that meet the World Health Organization’s (WHO) Target Product Profiles (TPPs) for triage (>90% sensitivity; >70% specificity) and detection (≥65% sensitivity; ≥98% specificity, with >98% sensitivity in culture positive, smear-positive patients) diagnostic tests ^6-9^. Here, we use the Diversity Outbred (DO) mouse population focusing on biomarker discovery to accelerate TB diagnostics. The benefit of the DO population is its genetic diversity (which is equivalent to human genetic diversity ^10^), and explains extreme responses to *Mtb* infection. Some DO mice are “supersusceptible” and develop early mortality within 8 weeks, due to lung damage, granuloma necrosis, inflammation, and weight loss ^11^. These human-like disease features and range of responses do not develop in the commonly-used C57BL/6 inbred strain, and rarely develop in other inbred mouse strains ^12,13^.

Here, we infected hundreds of DO mice with a low dose of aerosolized *Mtb* and examined gene expression profiles and inflammatory and immunological mediators for potential biomarkers. After exhaustively searching statistical models and machine learning classifiers to identify optimal sets of biomarkers, on independent testing, two classification algorithm/biomarker combinations achieved higher sensitivities and specificities than the WHO TPPs: Logistic Regression with matrix metalloproteinase 8 (MMP8) and Gradient Tree Boosting ^14^ with a panel of four biomarkers (CXCL1, CXCL2, tumor necrosis Factor (TNF), interleukin-10 (IL-10)). MMP8 and CXCL1 are significantly higher in human pulmonary TB patients than latent *Mtb* infection (LTBI) and normal controls. We show the DO population’s capacity to model human TB allows identification and validation of diagnostic biomarkers with strong translational potential to improve diagnosis of human pulmonary TB patients.

## Methods

### Ethics Statement

Tufts Institutional Animal Care and Use Committee (IACUC) and the Institutional Biosafety Committee (IBC) approved experiments under IACUC protocols: G2012-53; G2015-33; G2018-33 and IBC registrations GRIA04; GRIA10; and GRIA17.

### Mice and *Mtb* infection

Female DO and C57BL/6J mice were purchased from The Jackson Laboratory (Bar Harbor, ME) and housed in sterile BSL3 conditions at the New England Regional Biosafety Laboratory (Tufts University, Cummings School of Veterinary Medicine, North Grafton, MA). At 8-10-weeks old, mice were infected aerosolized *Mtb* strain Erdman bacilli (∼100 in experiments 1 and 2; ∼25 in experiments 3, 4, 5) using a CH Technologies system. We examined mice daily for weakness/lethargy, respiratory distress, and body condition score ^15^. Mice were weighed at least twice per week. All mice were confirmed to be infected by recovering live bacilli from homogenized lung tissue as described ^11,16^.

### Lung RNA expression profiling and analysis

One lung lobe was homogenized in TRIzol™ and stored at −80C until extraction using Pure Link RNA mini-kits (Life Technologies, Carlsbad, CA). RNA was checked for purity, and 42 lung RNA samples were analyzed at the Boston University Microarray and Sequencing Resource Core Facility (Boston, MA). Briefly, Mouse Gene 2.0 ST CEL files were normalized to produce gene-level expression values using the implementation of the Robust Multiarray Average (RMA) ^17^ in the affy package (version 1.62.0) ^18^ included in the Bioconductor software suite ^19^ and an Entrez Gene-specific probeset mapping (17.0.0) from the Molecular and Behavioral Neuroscience Institute (Brainarray) at the University of Michigan ^20,21^. Array quality was assessed by computing Relative Log Expression (RLE) and Normalized Unscaled Standard Error (NUSE) using the affyPLM package (version 1.59.0). The CEL files were also normalized using Expression Console (build 1.4.1.46) and the default probesets defined by Affymetrix to assess array quality using the AUC metric. Moderated t tests and ANOVAs were performed using the limma package (version 3.39.19) (i.e., creating simple linear models with lmFit, followed by empirical Bayesian adjustment with eBayes). Correction for multiple hypothesis testing was accomplished using the Benjamini-Hochberg false discovery rate (FDR) ^22^. Human homologs of mouse genes were identified using HomoloGene (version 68) ^23^. All microarray analyses were performed using the R environment for statistical computing (version 3.6.0). Enrichr (https://amp.pharm.mssm.edu/Enrichr) was used to determine the overrepresentation of Gene Ontology (GO) biological processes (version 2018) within an input set of official mouse gene symbols.

### Quantification of protein biomarkers in DO mouse lungs & human sera

Two lung lobes from each mouse were homogenized in 2mL of phosphate buffered saline and stored at −80C until use. Lung samples were serially diluted and tested for CXCL5, CXCL2, CXCL1, TNF, MMP8, S100A8, IFN-γ, IL12p40, IL-12p70 heterodimer, IL-10, and VEGF by sandwich ELISA using antibody pairs and standards from R&D Systems (Minneapolis, MN), Invitrogen (Carlsbad, CA), eBioscience (San Diego, CA), or BD Biosciences (San Jose, CA, USA), per kit instructions.

Serum samples were obtained from the Foundation for Innovative Diagnostics (FIND; Geneva, Switzerland). Patients were HIV-negative adults diagnosed with ATB by clinical signs and confirmed with sputum microscopy or culture. Patients diagnosed with LTBI did not have clinical, radiographical, or microbiological evidence of disease but were immunologically reactive to *Mtb* antigens. For additional details see Table S6. MMP8 and CXCL1 were quantified by ELISA using antibody pairs and standards from R&D Systems (Minneapolis, MN), per kit instructions.

### Statistical Analyses

Survival, weight, and ELISA data were analyzed and graphed in GraphPad Prism 8.4.2 with significance set at *p* < 0.05 and adjusted for multiple comparisons. Survival curves were analyzed using Log-rank (Mantel-Cox) test. For lung biomarkers and weights, data was analyzed for normal or lognormal distributions prior to Kruskal-Wallis one-way ANOVA and Dunnett’s post-tests, ***p<0.001. In DO mouse studies, sample sizes were determined based on a recent systematic review with recommendations for human biomarker studies ^6^. The studies using human sera were pilot studies to generate proof-of-concept results. All available human serum samples were used in ELISA testing. The AUC analysis on the human sera samples is done using GraphPad Prism 8.4.3. If there are multiple operating points that satisfies the >90% sensitivity and >70% specificity, we selected the one with the higher Youden’s index. If there none satisfy the criterion, we selected the operating point that’s closest to the region >90% sensitivity and >70% specificity.

### Machine learning algorithm development, implementation and validation

#### Discovery cohort

The lung protein measurements from 4 experimental infections with 107, 84, 60, and 231 samples respectively are combined into a dataset with 482 mice in total. Then, NI mice are removed, resulting in 453 mice. In order to handle missing data, we have considered two scenarios. In the first one, 407 mice which do not have any missing values in the following seven biomarkers CXCL5, CXCL2, CXCL1, IFN-γ, TNF, IL-12, IL-10, and the Lung *Mtb* burden measurement are selected. In the second scenario, 345 mice that do not have any missing lung biomarker values are selected. During feature selection if any of the three biomarkers MMP8, VEGF, S100A8 are included in a biomarker combination than the second set is used and if none of them are included then the first set is used. The samples are split 75% for classifier training and validation and 25% for classifier testing, stratified by class (Supersusceptible (SS), and not Supersusceptible (nSS)) and experiment number.

#### Leave-one-experiment-out setting

In the leave-one-experiment-out setting, three of the four experiments in the discovery cohort are combined and used as the training set and the remaining one is used as the validation set. Resulting in four different train/validation pairs. When the training portion of the discovery cohort is used the training and validation set sizes of the four pairs are 253,59; 267,45; 269,43;147,165 respectively and when the testing portion of the discovery cohort is used the training and validation set sizes of the four pairs are 253,17; 267,13; 269,12;147,53 respectively. If any of the biomarkers MMP8, VEGF, S100A8 are included in the subset selection then only the samples with complete measurements for all ten biomarkers are used.

#### Independent cohort

In the independent cohort, there are 122 samples (93 nSS, 23 SS, 6 Control) and there weren’t any samples with missing values.

#### Experiment-wise metrics

To calculate the experiment-wise sensitivity (specificity), first, the sensitivity (specificity) in each experiment is calculated and then averaged. Minimum experiment sensitivity (specificity) is calculated by first obtaining the sensitivity (specificity) in each of the experiments and then taking the minimum. In the leave-one-experiment-out setting, experiment-wise sensitivity (specificity) is defined as the average of the four sensitivity (specificity) values. Similarly, minimum experiment sensitivity (specificity) is the minimum of the four sensitivity (specificity) values. To calculate the AUC, an experiment-wise Receiver Operating Characteristics (ROC) curve is drawn where the y-axis corresponds to sensitivity and the x-axis to the specificity and then the area under that curve is calculated.

#### Preprocessing

Each biomarker is standardized by subtracting the sample mean and dividing it by the uncorrected sample standard deviation. During training, only training samples are used to estimate the population parameters. For the first approach during testing, both the samples of the training and the testing sets are used to estimate the population parameters to standardize the testing samples. This holds for testing on Discovery Cohort and Independent Cohort as well. For the second approach, the population parameters estimated in training are used to standardize the test set.

#### Classification algorithm, hyper-parameter and feature selection

We have used two different variations of best-subset selection. Using the first approach, we have identified the Logistic Regression with MMP8 and using the second approach we have identified the Gradient Tree Boosting with CXCL1, CXCL2, TNF, and IL-10. During both of the approaches, for each subset of features in the search space, all classifiers and their hyper-parameters are searched. For each subset and each classifier, hyper-parameters with the highest AUC are selected for the first approach, and for the second approach, the ones with the highest experiment-wise AUC are selected. In order to evaluate the out-of-sample performance 5-fold Cross-Validation (CV) and 100-fold CV for the first and the second approaches respectively.

#### Subsets searched

The feature search space has all the combinations of the CXCL5, CXCL2, CXCL, IFN-γ, TNF, IL-12, IL-10, and in addition to them all combinations of the CXCL2, CXCL1, IL-10, IL-12, MMP8, VEGF, S100A8 are searched, resulting in 239 different selections.

#### Classification algorithms searched

We have compared the performance of linear classifiers Support Vector Machine (SVM), Logistic Regression with L1 regularization and non-linear classifiers SVM with Radial Basis Function, Random Forest, and Gradient Tree Boosting through 5-fold-CV in the training portion of the discovery cohort. Observing the performance between linear classifiers and non-linear classifiers were similar in their respective categories, we have selected one from each category. The selected two classifiers were Logistic Regression with L1 regularization and Gradient Tree Boosting. Scikit-learn ^24^ is used for the implementation of the classifiers. The number of samples in each class is unbalanced; therefore, each sample is re-weighted by its inverse class proportions in the loss functions of both classifiers.

#### Hyper-parameters searched

For Gradient Boosting Tree, logloss is used and as hyper-parameters learning rate (0.001, 0.01), number of trees (1,3,5), and max depth of a tree (1,3,5) are searched for the first approach. For the second approach (0.01, 0.3, 0.5,1.) are searched for the learning rate instead. For the Logistic Regression with L1 regularization, the weight of the L1 loss is selected among (0.01,0.0316, 0.1, 0.316, 0.1) for both approaches. As the result of the search, for the biomarker panel of CXL1, CXCL2, TBF and IL10, learning rate 0.1, number of trees 3, and max depth of a tree 5 are selected. For the Logistic Regression with MMP8 all hyper-parameters we have observed that the weight of L1 loss didn’t change the AUC for a classifier with a single feature.

#### Retraining after classifier selection

After the classifier selection is complete, the selected classifier with fixed hyper-parameters and selected features is re-trained on all training portion of Discovery Cohort, then evaluated on the testing portion of it. Similarly, the selected classifier is trained on all of the Discovery Cohort before it’s evaluated on the independent cohort.

#### Operating point selection

We have selected the prediction threshold as a hyper-parameter and selected it through K-fold-CV. In the first approach, 5-fold-CV is used and the threshold that achieves the highest sensitivity while maintaining at least 70% specificity is selected. In the second approach, in order to reduce the error resulting from selecting a single threshold for K-classifiers we have used a higher number of folds where during the classifier selection 100-fold-CV is used and 150-fold-CV is used when the selected classifier is retrained using both training and the testing datasets. The operating point that maximizes the experiment-wise sensitivity while achieving at least 70% specificity in each of the experiments is selected. If two operating points satisfy the criteria and achieve a similar (<0.1%) experiment-wise sensitivity the one with the higher experiment-wise specificity is selected.

## Results

### Mtb infection and pulmonary TB in DO mice

One third of DO mice are supersusceptible (SS) to *Mtb*, developing early-onset disease within 8 weeks of aerosol infection by ∼25 bacilli, significantly reducing survival compared to age- and sex-matched non-infected DO mice or to *Mtb-*infected C57BL/6J inbred mice (Figure 1A). The SS phenotype is consistent across sexes, institutions, and aerosol infection methods ^11,25^. Peak mortality occurs 20-35 days after *Mtb* (Figure 1B) and disease onset begins as early as 4 days after infection (Figure 1C). The remaining ∼70% of *Mtb-*infected DO mice and all C57BL/6J mice show no morbidity or mortality over 8 weeks.

**Figure 1.**
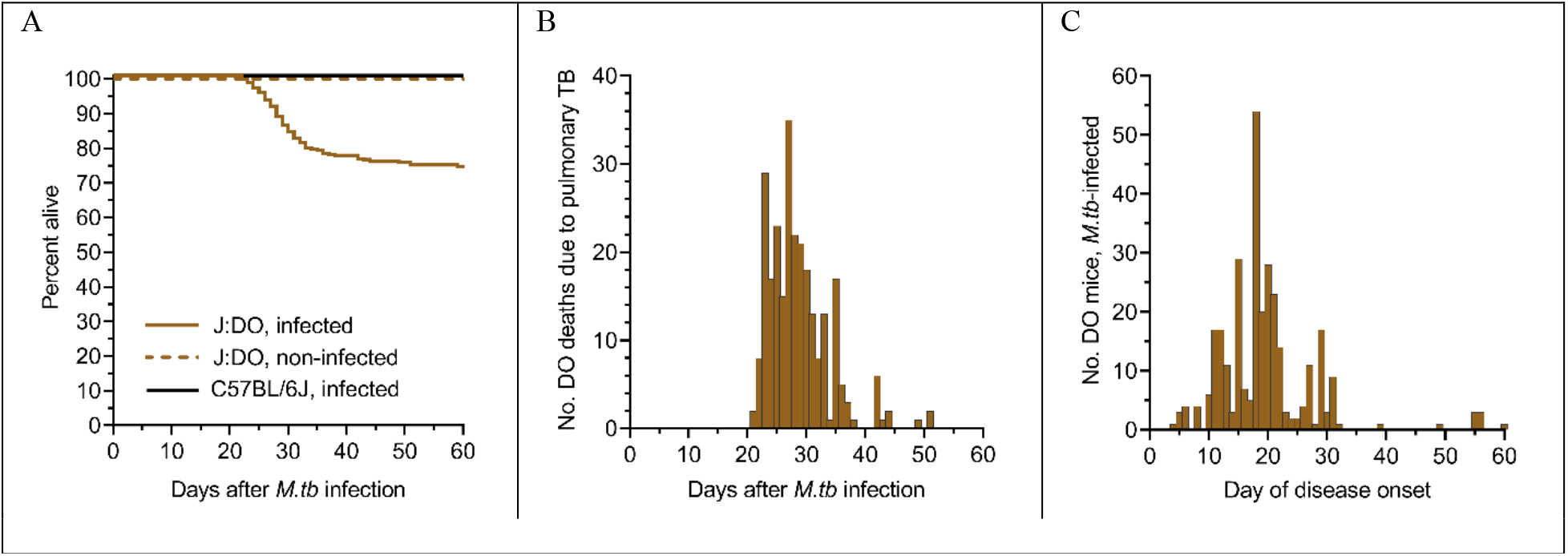
Survival, mortality, and disease onset. We infected 8-10-week-old, female DO (n=657) and C57BL/6J (n=66) mice with ∼25 *Mtb* bacilli by inhalation, monitored, and euthanized if morbidity occurred. Within 8 weeks, 34% of DO mice succumned to pulmonary TB, while 66% survived. All non-infected DO mice (n=40) and all *Mtb-*infected C57BL/6J inbred mice survived without morbidity or mortality for at least 8 weeks (*p* < 0.0001 by Log-rank Mantel-Cox test) (A). Deaths in SS DO mice occurred between 21-52 days and peaked 25-35 days (B). Disease onset in SS DO mice began as early as 4 days after *Mtb* infection and peaked at 18 days post-infection (C).

### Lung gene expression identifies inflammatory mediators as potential biomarkers of supersusceptibility

Immune deficiency does not explain early-onset TB in the DO population. DO mice generate *Mtb* antigen specific TH1 immunity, respond to *M. bovis* BCG vaccination without developing BCGosis ^11,16,25^ and generate TH2 and TH17 immune mediators (Supplemental Figures). Our prior work identified three neutrophil chemokines (CXCL1, CXCL2, CXCL5) that diagnosed SS DO mice with modest accuracy ^11^. To identify more biomarkers and gain mechanistic insight, we followed 32 *Mtb-*infected DO mice for 157 days, and profiled total lung RNA for gene expression by microarray. Ten mice developed pulmonary TB before 8 weeks and 22 *Mtb-*infected DO mice showed no evidence of disease. Five age- and sex-matched non-infected DO controls were also profiled. An analysis of variance revealed that 20,623 of the 25,206 interrogated genes were significantly differentially expressed (FDR *q* < 0.05) across the three groups. Additional filters (fold change >2) identified 121 genes specifically and highly expressed in lungs of SS DO mice (Figure 2). Of those 121 genes, three: *S100a8, Mmp8*, and *Cxcl2* appeared in 18 of 31 pathways identified using Enrichr (Table S1). Mechanistically, all gene pathways specific to SS converge on acute inflammatory responses: Neutrophil (granulocyte) recruitment and degranulation; positive inflammatory signaling, cytokines and chemokines; and extracellular matrix degradation.

**Figure 2.**
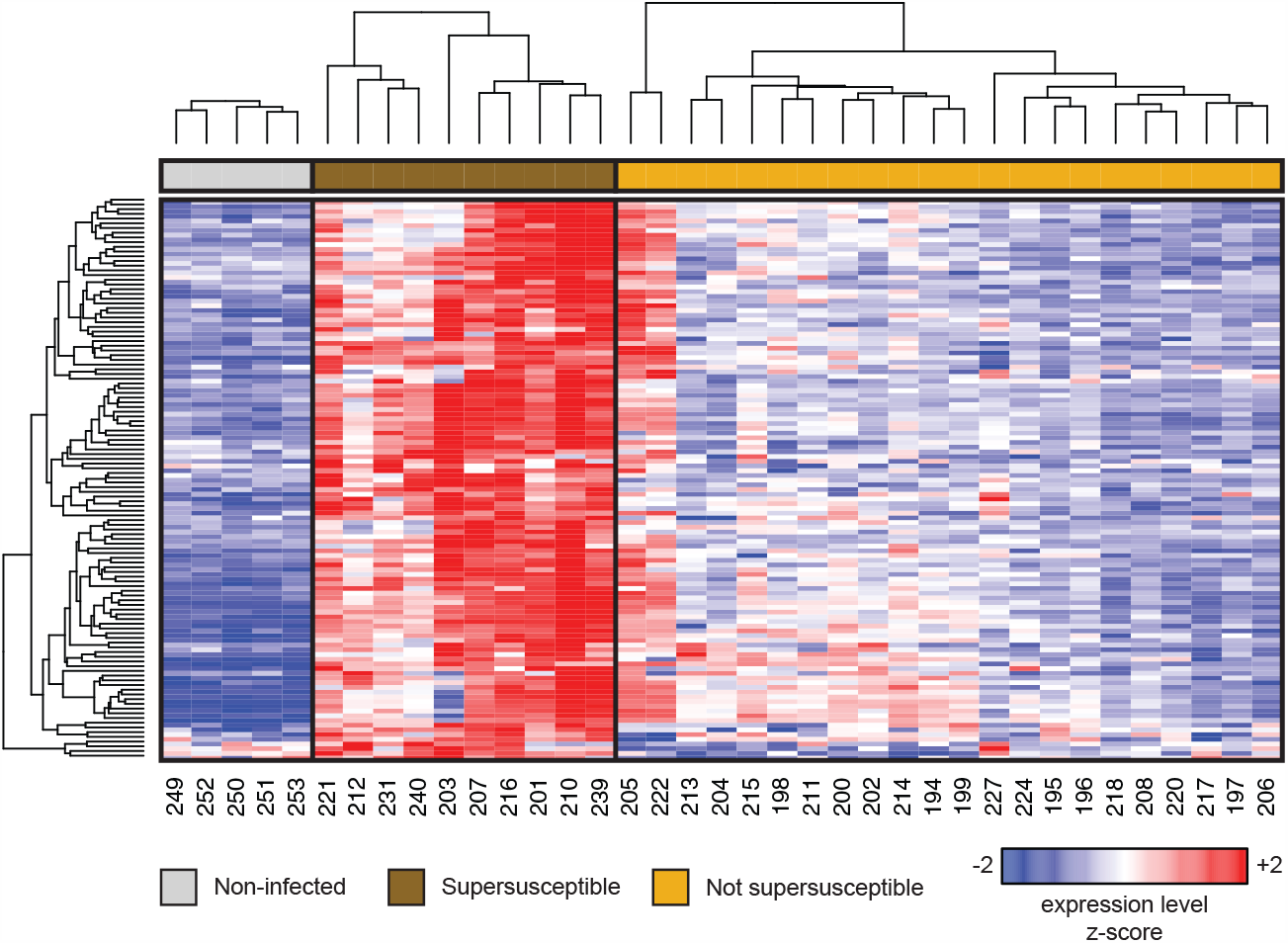
121 genes are unniquely and highly expressed in lungs of SS DO mice, compared to other groups. Microarray gene expression profiling identified a set of 121 genes that changed significantly >2-fold in SS (dark brown) relative to non-infected (gray) and to not-supersusceptible (tan) DO mice. Rows and columns correspond to genes and individual DO mice, respectively. Hierarchical clustering was performed across all rows and was also performed separately within each group. We z-normalized the expression values for each gene to a mean of zero and standard deviation of one across all samples in each row; blue, white, and red indicate z-scores of ≤ −2, 0, and ≥ +2, respectively.

### Immune and inflammatory proteins in lungs of Mtb-infected mice

To generate data sets for classifier and biomarker selection, we used CXCL2, S100A8, and MMP8 proteins (products of highly expressed genes in SS DO mouse lungs identified by microarray); TH1 proinflammatory cytokines required for resistance to *Mtb* (IFN-γ, TNF, and IL-12 (p40 and p70); immune suppressive IL-10; and neutrophil chemokines that cause inflammation in *Mtb* infected lungs: CXCL1, CXCL5 ^26-28^. We included Vascular Endothelial Growth Factor (VEGF) because VEGF is a component of a promising biomarker panel ^7^. We measured proteins in the lungs of *Mtb-*infected DO mice, C57BL/6J mice, and non-infected age- and sex-matched control DO mice (Figure 3, panels A-K). All biomarkers except IL-12p40, IL-12p70 heterodimer, and VEGF were significantly higher in the lungs of SS DO mice compared to all other groups.

**Figure 3.**
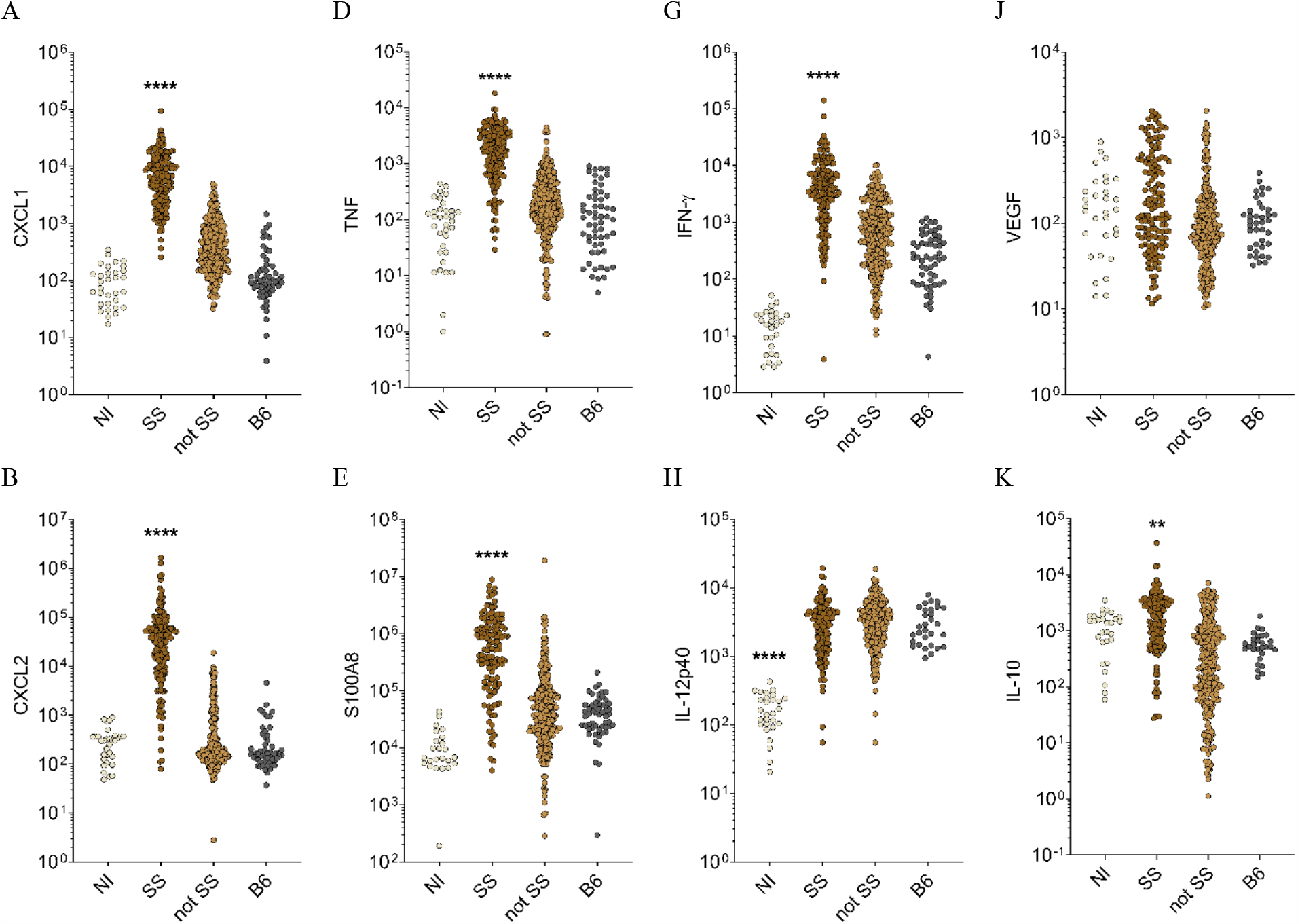
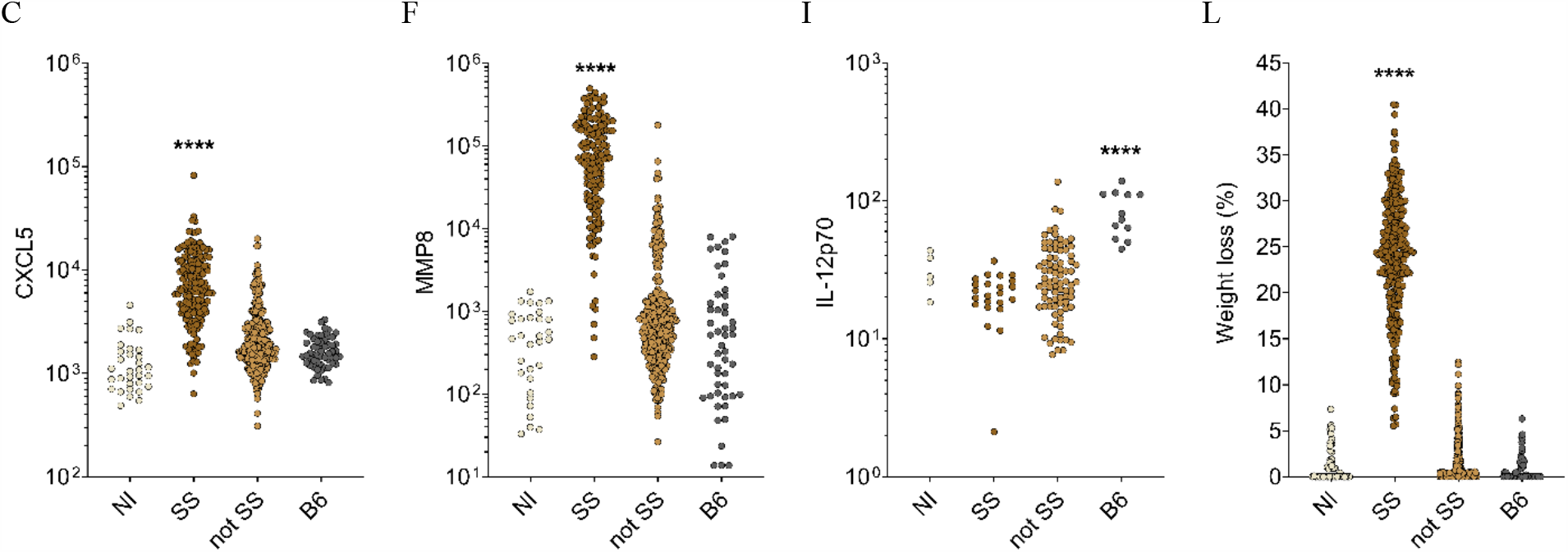
Lung biomarkers and morbidity in *Mtb-*infected mice. We infected 8-10-week-old, female DO and C57BL/6J mice with ∼25 *Mtb* bacilli by inhalation, and euthanized 8 weeks later or sooner if morbidity developed. All supersusceptible (SS) DO mice succumed within 8 weeks, while the rest showed no morbidity or mortality. We measured lung biomarkers using commercial sandwich ELISAs (A-K). At euthanasia, weight loss was calculated for each mouse as a percent compared to its maximum (L). All data were lognormal distributed and analyzed by Kruskal-Wallis one-way ANOVA with Dunnett’s multiple comparisons post-tests (**p<0.01; ****p<0.0001). Each dot repsesents 1 mouse. Results shown are combined from 5 independent experimental infections.

### Two different biomarker panels are identified using machine learning

To identify diagnostic biomarkers, we organized data from five different experimental infections into discovery and independent cohorts (Figure 4).

**Figure 4.**
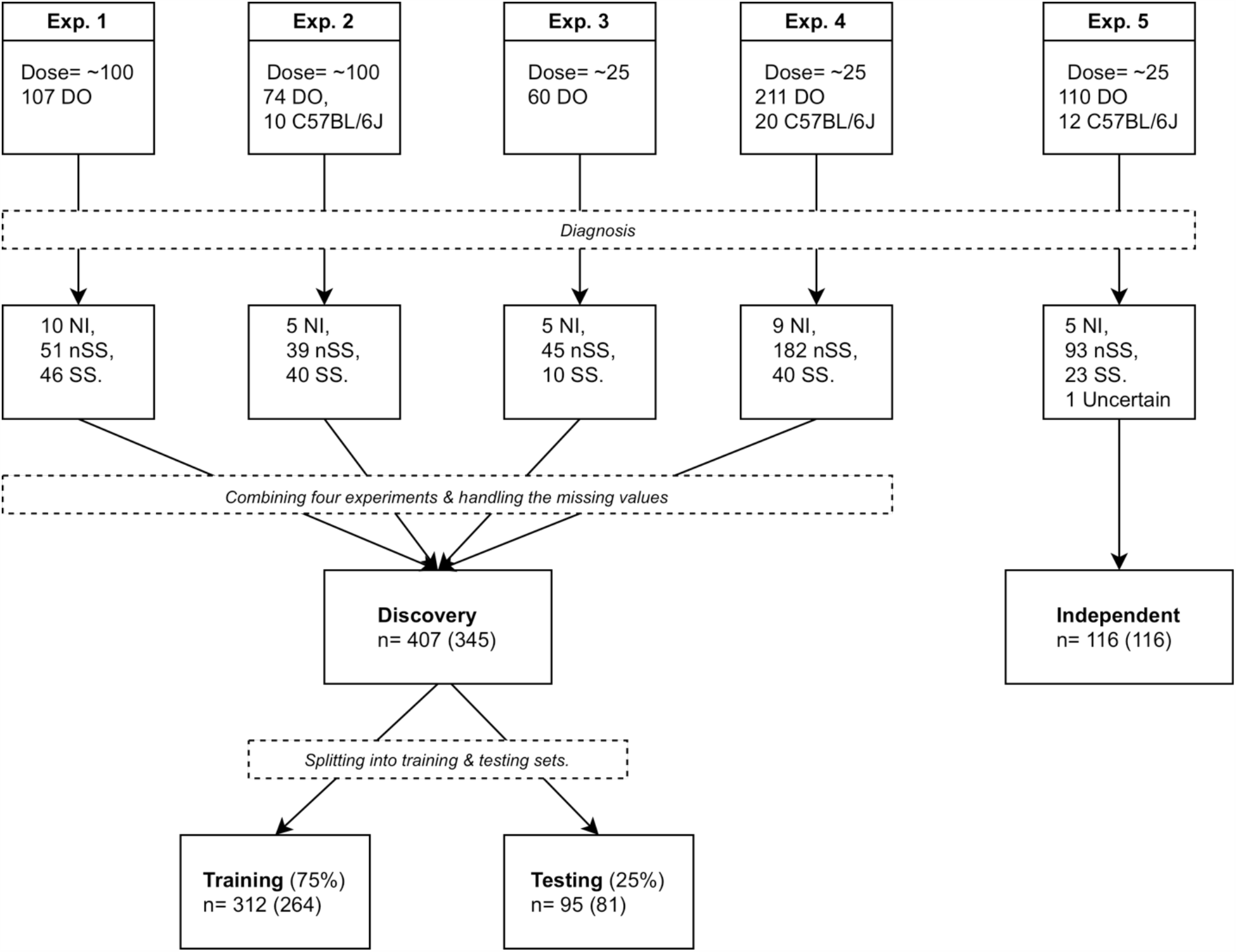
Flow chart of sample organization and datasets. The top most five boxes denote the initial *Mtb* dose and the number of DO and C57BL/6J (if used) mice for each of the experiments. The succeeding five boxes report the number of mice in each disease class. We used the nSS and SS mice from Exp. 1, Exp. 2, Exp. 3 and Exp. 4 in the discovery phase and nSS and SS mice from Exp. 5 in the independent evaluation. The first number to the right of “n=” denotes the number of samples that do not have any missing values in CXCL5, CXCL2, CXCL1, IFN-γ, TNF, IL-12, IL-10, and the Lung Mtb burden measurements, and the second one within the paranthesis denotes the number of samples that do not have any missing lung biomarkers.

First, we applied best-subset feature selection with 5-fold cross validation to the training data. From the 478 classifiers (a classifier refers to a specific classification algorithm and specific biomarkers) Logistic Regression with MMP8 (Table S2) achieved 0.95 AUC, 94.1% sensitivity and 87.4% specify for classifying SS and nSS mice. To avoid overfitting, we validated performance of Logistic Regression with MMP8 on the test data, achieving 0.96 AUC with 94.4% sensitivity and 87.3% specificity. On data from the independent cohort, the classifier achieved 0.987 AUC (Figure S1A), 78.3% sensitivity, 100% specificity, 100% positive predictive value (PPV) and 94.9% negative predictive value (NPV). Although its sensitivity was below the TPP triage test target, Logistic Regression with MMP8 was highly attractive because it used a single biomarker and a linear decision boundary. However, the classifier had unacceptable variation in specificity across experiments, which led us to investigate whether we could identify a different classifier that performed with greater consistency.

We sought a classifier capable of 90% sensitivity and 70% specificity in each experimental infection used in discovery. Among the identified classifiers only three (Table S3) had >80% minimum experiment sensitivity and 70% minimum experiment specificity in the leave-one-experiment-out setting. From the three, we pursued Gradient Tree Boosting with the biomarker panel CXCL1, CXCL2, TNF, and IL-10 because it achieved the highest minimum experiment sensitivity (93.3%), had high experiment-wise sensitivity (97.3%) and overall sensitivity (96.6%). In the testing portion of the discovery cohort, the four-biomarker panel (CXCL1, CXCL2,

TNF, and IL-10) achieved 96.9% and 94.4% sensitivity and specificity, respectively, across all four experiments combined. In the leave-one-experiment-out setting, this classifier had 87.5% and 70% minimum experiment sensitivity and specificity, respectively. In the independent cohort, the four-biomarker panel achieved 91.3% sensitivity and 81.7% specificity, satisfying the WHO performance criteria for a triage test. The PPV and NPV of the classifier were 55.3% and 97.4%, respectively, and its AUC was 0.95 (Figure S1B). We evaluated the classifier further using the 27 age- and sex-matched non-infected DO controls and it achieved 100% specificity. Of the 4 biomarkers in the panel, CXCL1 and CXCL2 were more important than TNF and IL-10, shown by higher variable importance values of 0.62, 0.32, 0.04, and 0.02, respectively.

To determine how each biomarker contributes to classifier performance, we measured the average percent difference. MMP8 achieved the highest effect, with an average percent difference of 11.31%, followed by CXCL1 (5.8%) and CXCL2 (4.82%) (Table S4). Figure 5 demonstrates this effect graphically by showing that biomarker panels lacking CXCL1, CXCL2, or MMP8 have lower AUC values than the biomarker sets that contain at least 1 of these 3.

**Figure 5.**
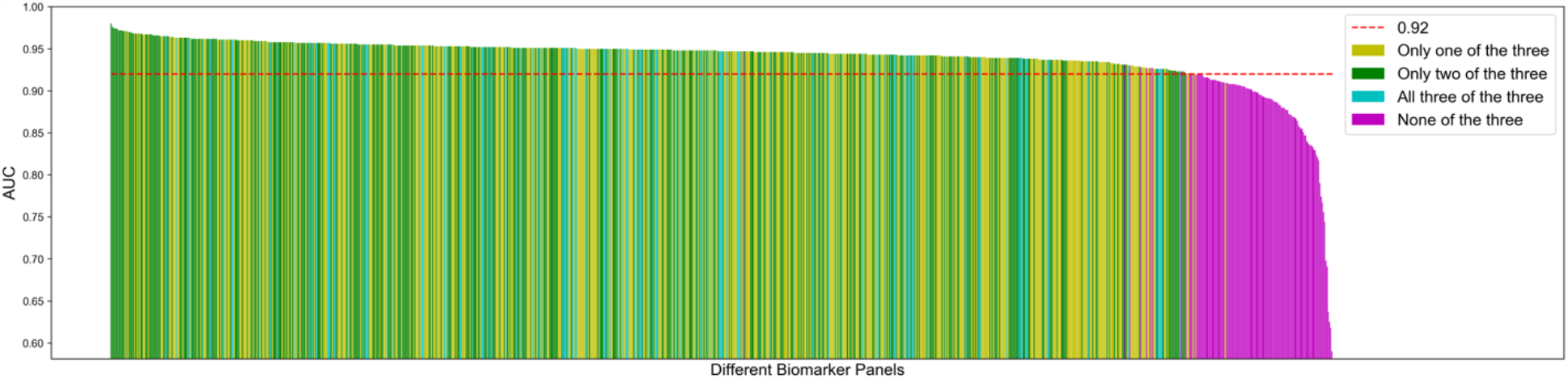
Classifiers using lung protein biomarkers CXCL1, CXCL2, or MMP8 have the highest performance. Bar chart of 5-fold-cross validation AUC of 1023 different biomarker panels sorted in descending order. Y-axis denotes the AUC and each bar in the x-axis corresponds to a different panel. Yellow, green and teal bars indicate the classifiers that are using any one of the three biomarkers, any two of three biomarkers, and all three biomarkers respectively. Magenta bars indicate the classifiers that did not include CXCL1, CXCL2 or MMP8. Biomarker panels that did not include CXCL1, CXCL2, or MMP8 had lower AUC, shown by the red dashed line, at 0.92.

### MMP8 and CXCL1 biomarkers have translational potential for human pulmonary TB

For human study, we pursued MMP8 and CXCL1 because neither have been fully investigated in humans and both performed well under different conditions in DO mice. MMP8 is the biomarker used in the first classifier and CXCL1 had the highest variable importance among the panel used in the second classifier. We obtained human sera from HIV-negative adults with active pulmonary TB (ATB) or LTBI from the Foundation for Innovative Diagnostics. Pooled serum from healthy individuals served as controls. MMP8 and CXCL1 protein levels were significantly higher in sera from patients with ATB to other groups (Figure 6). For ATB vs normal sera, MMP8 had perfect AUC (1.00), 100% sensitivity and specificity; and CXCL1 had 0.940 (0.892 to 0.989) AUC, 91.0% (81.8% to 95.8%) sensitivity and 90.9% (72.2% to 98.4%) specificity. For ATB vs LTBI, MMP8 had 0.676 (0.578 to 0.773) AUC, 60.6% (48.5% to 71.5%) sensitivity and 64.6% (50.4% to 76.6%) specificity; and CXCL1 had 0.844 (0.771 to 0.918) AUC, 83.6% (72.9% to 90.6%) sensitivity and 66.7% (52.5% to 78.3%) specificity.

**Figure 6.**
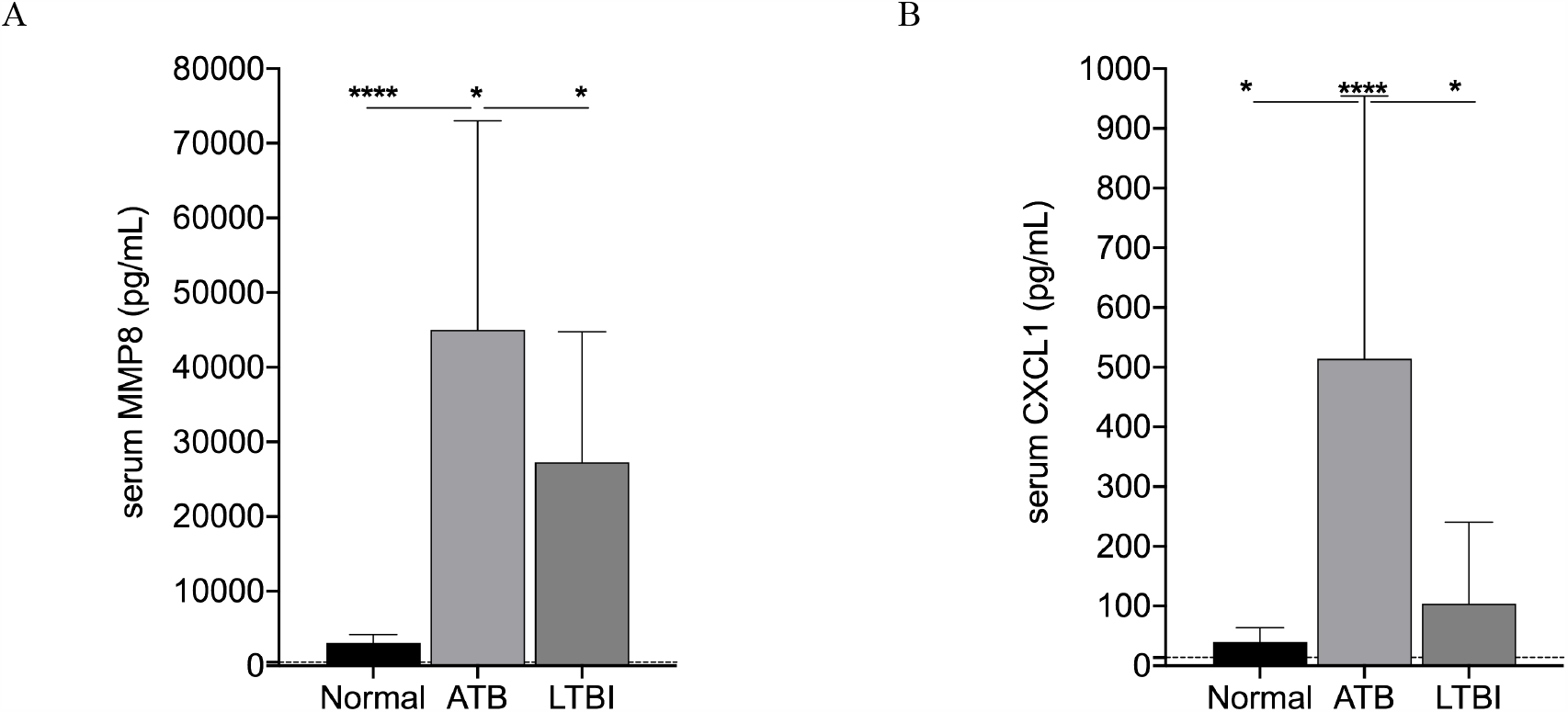
MMP8 and CXCL1 in human sera from patients with active pulmonary TB (ATB), latent *Mtb* infection (LTBI), and normal individuals. We tested serum from HIV-negative patients ATB (n= 66 and 67 for MMP8 and CXCL1 respectively), LTBI (n=48) and replicates from pooled normal (n=24) for MMP8 (A) and CXCL1 (B) by ELISA, and analyzed data by Kruskal-Wallis one-way ANOVA with Dunnett’s multiple comparisons post-tests (*<0.05, ****p<0.0001). Dashed lines show the limit of detection (LOD): MMP8 LOD = 480.7pg/mL and CXCL1 LOD = 13.89 pg/mL

## Discussion

WHO introduced the TPPs in 2014, to unify and promote development efforts for TB diagnostics in the direction of end-user requirements ^9^. A systematic review of biomarker papers between 2010 and 2017 ^6^, identified 44 panels or single biomarkers that satisfies a TPP criterion. However, in terms of the QUADAS-2 assessments (study design, sampling, negative population, timing, reference standard and blinding), only one panel had a low risk of bias ^6^. The panel included ECM1, myoglobulin, HCC1, IL-21, ENA-78, TPA, IL-12(p40) and IL-13 which achieved 100% (95% CI, 83.2–100%) sensitivity and 95% (95% CI, 68.1–99.9%) specificity with a small sample size n=39 ^29^. A more recent study, identified a panel of IL-6, IL-8, IL-18, VEGF, and anti-Ag85 which performed well, only slightly below the TPP triage test (86% sensitivity and 69% specify) on hundreds of samples from geographically distinct regions ^7^. Guidance from the systematic review and results from Ahmed *et al* demonstrate a clear need for further biomarker discovery, validation, and independent testing with minimal bias, on large numbers of samples.

To address that need and accelerate TB biomarker discovery, we modeled human responses to *Mtb* by using the DO mouse population. Like humans, this population is highly genetically diverse and demonstrates variable responses and disease outcomes to *Mtb*, ranging from supersusceptible to highly resistant ^11,25^. We classify *Mtb-*infected DO mice as supersusceptible based on survival <8 weeks. Our supersusceptible DO class is somewhat analogous to progressor DO mice recently described ^30^. We prefer survival for ground truth as the early peak in DO mortality is robust and reproducible across hundreds of mice, while *Mtb* burden and inflammation scores (used to identify DO progressors) fall along a continuous spectrum and become unstable ground truth as mouse numbers increase. Regardless, the richness of the DO responses to *Mtb* (Figures 3, 4) clearly provides an unprecedented tool for discovery, validation, and testing on independent cohorts.

We identified two classifiers that can accurately discriminate between supersusceptible and not-supersusceptible, namely Logistic Regression with MMP8 and Gradient Tree Boosting with a panel of four biomarkers (CXCL1, CXCL2, TNF, and IL10). The first classifier had an advantage of being a single biomarker and a simpler regression model, with MMP8 satisfying the minimal requirements of a WHO TPP detection test. However, its performance showed batch variation across experiments attributed to the different criterion used in feature selection. The second classifier had an advantage of performing consistently across experimental batches. Gradient Tree Boosting with CXCL1, CXCL2, TNF, and IL10 also achieved the WHO triage test specifications with the operating points selected in the discovery cohort.

To our knowledge, using DO mice for translational biomarker discovery has not been explored previously. Two highly performing biomarkers in DO mice were MMP8, the only biomarker used in the first classifier, and CXCL1, the biomarker that achieved the highest variable importance in the second classifier. Additionally, expression of Mmp8 gene was significantly higher in the lungs of supersusceptible DO mice compared to other groups. In human patients, both MMP8 and CXCL1 were significantly higher in patients active pulmonary TB compared to patients with LTBI or to pooled sera from normal individuals. In our proof-of-concept study, each biomarker had sensitivity and specificity values higher than the minimum requirements of the TPP triage test to distinguish ATB from pooled normal samples. Discriminating ATB from LTBI in human sera using MMP8 or CXCL1 was more challenging, likely related to the small sample size. The AUC of MMP8 has dropped to 0.676 but CXCL1 maintained a relatively high AUC (0.844) and its performance was only slightly below the triage test target.

MMP8 and CXCL1 have not been fully investigated in humans, and we suggest they be rigorously pursued. MMP8 and CXCL1 were not among the TB biomarkers with the highest AUC values reported in humans which were C-reactive protein, transferrin, IFN-γ, interferon (IFN)-γ inducible protein-10 (IP-10), IL-27, and interferon–inducible T Cell Alpha Chemoattractant (ITAC-1) ^31-39^. Only one study reported MMP8 performance as a biomarker in children (<25 per diagnostic category) ^40^ and three studies measured CXCL1 ^41-43^. A few investigators quantified MMP2, MMP3 and MMP9 (proteins functionally related to MMP8) but did not pursue them further due to missing data ^7^, or inconsistent statistical differences ^31,39^. Three studies measured CXCL1 ^41-43^ comparing TB patients to non-TB patients, and further investigation is crucial because small sample sizes and variable sample types preclude definitive interpretation: n=44 to 88 for plasma ^41^; n=11 to 27 for serum ^42^; and n=11 to 27 and n=32 to 72 for saliva ^42,43^.

Our work here discovering, validating, and independently testing TB biomarkers in DO mice is important, yet has three limitations. First, we used two separate datasets in the discovery cohort (n=407 or 345) because of missing data. Ideally, when comparing two different classifiers, we should use identical data sets to estimate out-of-sample performance. However, since mouse lungs are small with limited volumes, not all samples were available to measure all biomarkers. This contributed to missing values, and for simplicity, samples with missing values were excluded from training, validation, and testing. Second, the WHO Target Product Profiles (TPPs) were intended for non-sputum samples of humans ^9^ and not lung samples from experimentally infected mice. We believe, however, that benchmarking against the TPPs increases the translational relevance of our findings and future studies will use non-lung samples from *Mtb-*infected DO mice. Third, the number of human serum samples we used is smaller than the recent biomarker publications such as ^7^ which has 179 TB and 138 other TB-like disease patients. Our aim, however, is to demonstrate a proof of concept that DO mice can discover translationally relevant TB biomarkers. We acknowledge that larger studies including TB patients with different forms of disease, co-morbidities and including patients with non-TB lung disease are required for final validation and independent testing in humans.

Overall, we show two important findings: First, that DO responses to *Mtb* exceed the ranges of inbred mice, clearly demonstrating genetic control of variable susceptibility to *Mtb*. Their responses mimics human disease and resistance phenotypes that are not observed in inbred mice, providing the field with a powerful *in vivo* model for mechanistic and biomarker discovery. Second, that MMP8 and CXCL1 identified, validated, and independently tested in lung samples from DO mice, are translationally relevant biomarker candidates for human TB that may improve our ability to diagnose TB patients, and are worthy of continued pursuit.

## Data Availability

The data is not currently publicly available.

## Contributors

DK – Methodology, formal analysis, data analysis, writing, review, editing the manuscript.

MKKN – Methodology, formal analysis, data analysis, writing and editing the manuscript.

TT – Methodology, formal analysis, data analysis, writing the manuscript.

MG – Sample collection, sample curations, sample quality control, assay performance, data generation.

YL – Sample testing and data analysis CM – Data analysis

AG – Data curation, quality control, analysis

DMG – Data curation, quality control, analysis.

IK – Funding acquisition, conceptualization, review, editing of the manuscript

MG – Funding acquisition, methodology, project administration, and review and editing of the manuscript.

BY – Funding acquisition, methodology, project administration, and review and editing of the manuscript.

GB – Funding acquisition, conceptualization, project administration; sample and data collection, assay performance, curation, data analysis, writing, review, editing of the manuscript.

## Acknowledgments

We thank Julie Tzipori, Curtis Rich, Donald Girouard, and Sam Telford III for services in the New England Regional Biosafety Laboratory at Tufts University Cummings School of Veterinary Medicine, North Grafton, MA. Frances Brown, Linda Wrijil, Sarah Ducat, and Gina Scarglia provided histology services at Tufts University’s Cummings School of Veterinary Medicine. All microarray protocols were carried out by the Boston University Microarray and Sequencing Resource (BUMSR) core facility, and we thank Eduard Drizik of the BUMSR for the initial analysis of this experiment. Support was provided by NIH R21 AI115038 (GB); NIH R01 HL145411 (GB); NIH UL1-TR001430 (BUMSR); and the American Lung Association Biomedical Research Grant RG-349504 (GB).

## Supplementary Materials

**Fig. S1.**
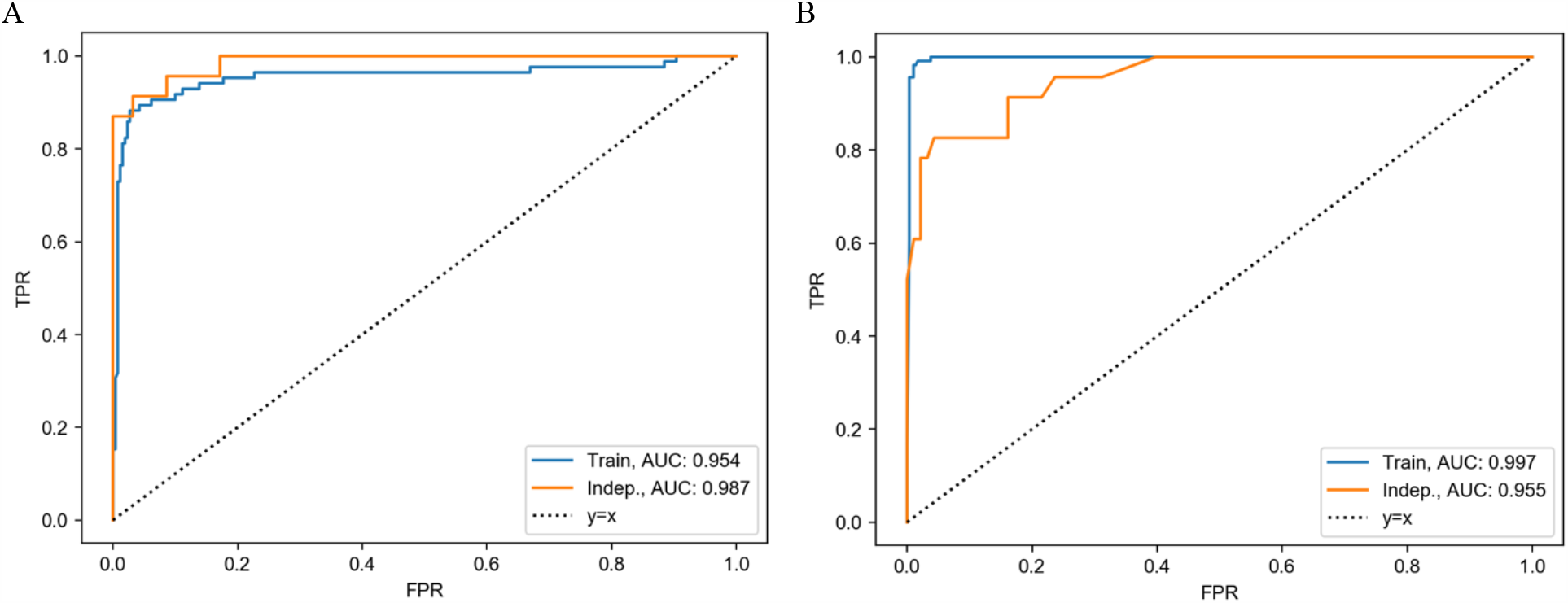
Receiver Operating Characteristics (ROC) curves of classifier performance to diagnose supersusceptible *Mtb*-infected DO mice. A) Logistic Regression with MMP8, and B) Gradient Tree Boosting with CXCL1, CXCL2, TNF, and IL-10. Blue and orange curves correspond to the AUC in the discovery cohort (n=345 for A and n=407 for B) and independent cohort (n=116) respectively for both A and B.

**Fig S2.**
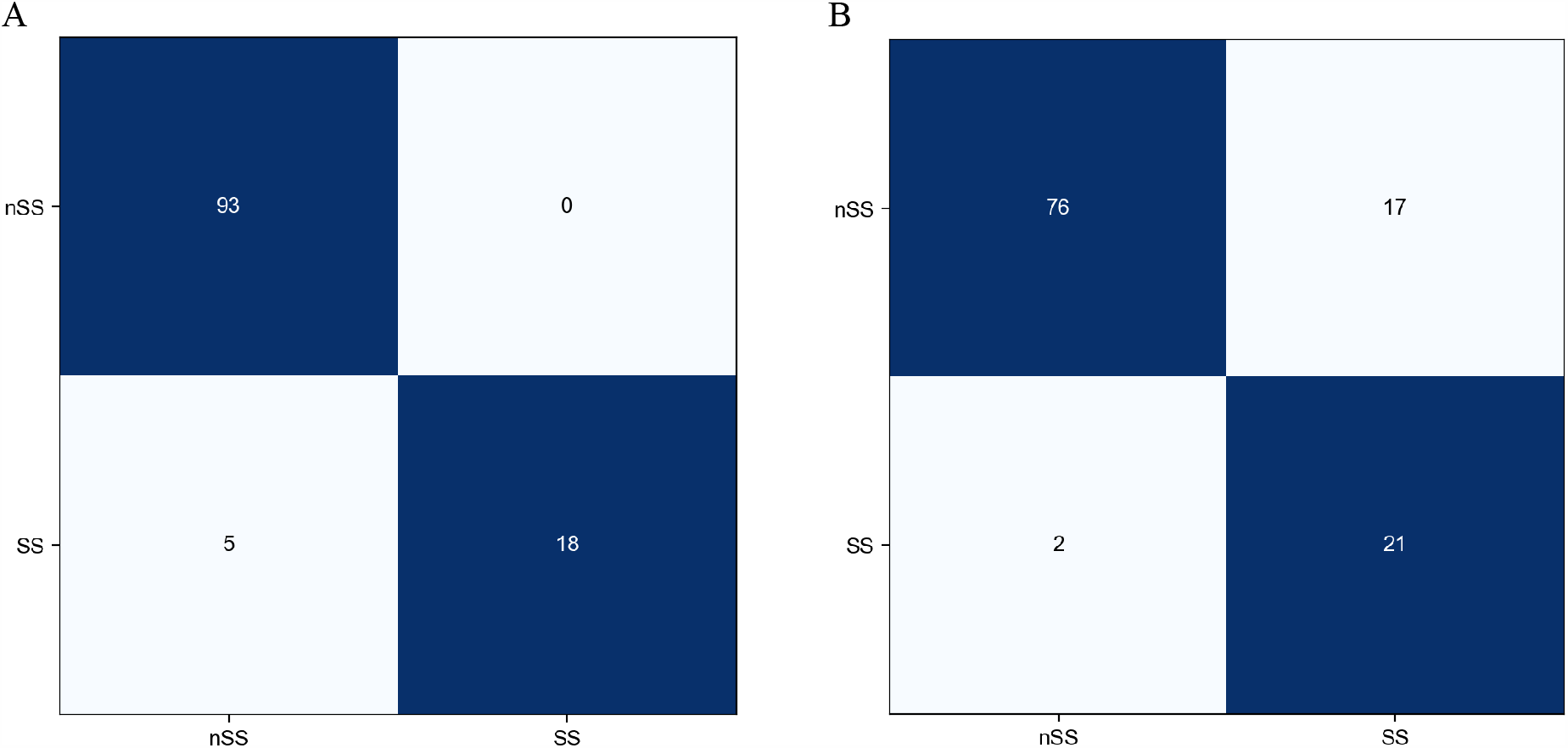
Confusion matrices for classifying between SS and nSS mice in the independent cohort. A) Logistic Regression with MMP8 and B) Gradient Tree Boosting with CXCL1, CXCL2, TNF, and IL-10. Row labels and column labels indicate the correct and the predicted labels respectively.

**Figure S3.**
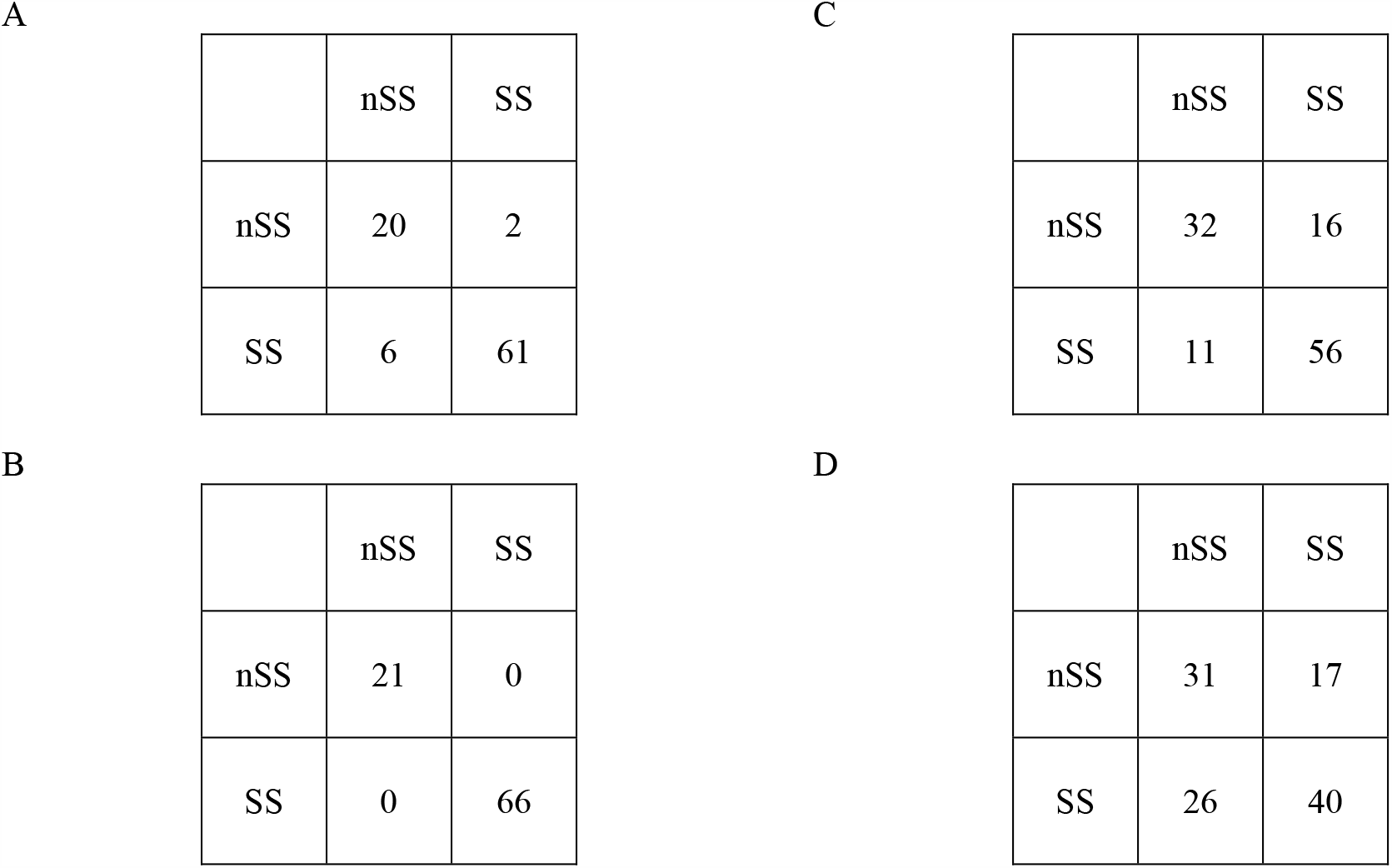
Confusion matrices for the biomarkers in human sera. A) and B) confusion matrices of CXCL1 and MMP8 for classifying between ATB and pooled normal samples. C) and D) confusion matrices of CXCL1 and MMP8 for classifying between ATB and LTBI. For each confusion matrix row labels and column labels indicate the correct and the predicted labels respectively.

**Fig. S4.**
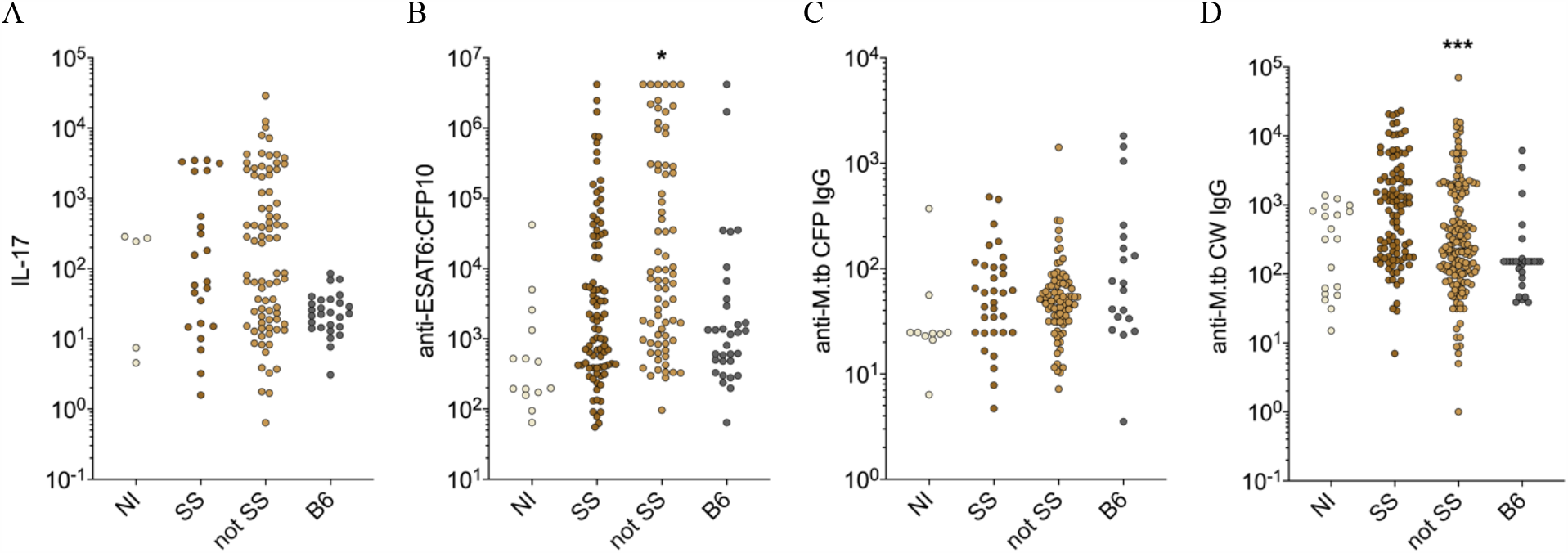
Additional lung biomarkers we measured are displayed in (A-D). All data were lognormal distributed and analyzed by Kruskal-Wallis one-way ANOVA with Dunnett’s multiple comparisons post-tests (*p<0.05; ***p<0.001). Each dot repsesents 1 mouse.

**Table S1.** Pulmonary TB in SS DO mice reflects acute inflammation, neutrophil recruitment/activation and extracellular matrix degradation. Enrichr identified the biological pathways within a set of 121 genes that were highly expressed in SS mice relative to non-infected DO mice and non-SS mice. GO terms with adjusted p < 0.01 are shown, along with the human homologs of the genes overlapping with the term. Expressed genes in bold were pursued as diagnostic biomarkers.

| Gene Ontology Term | GO Term ID | p | Adjusted p | Genes |
| --- | --- | --- | --- | --- |
| cellular response to cytokine stimulus | GO:0071345 | 1.82E-13 | 9.28E-10 | CCR1, IL11, IL1RN, CSF3R, IL1R2, FPR1, OSM, F13A1, ACOD1, IL1RAP, **CXCL2**, MMP9, EREG, IL1A, IL6, IL1B, CCL4, CCL3, SAA1, PRTN3, TIMP1 |
| cytokine-mediated signaling pathway | GO:0019221 | 1.90E-13 | 4.86E-10 | CCR1, IL11, IL1RN, CSF3R, SERPINB2, RSAD2, IL1R2, FPR1, OSM, F13A1, IL1RAP, PPBP, CXCL3, **CXCL2**, MMP9, EREG, IL1A, IL6, IL1B, CCL4, CCL3, SAA1, PRTN3, TIMP1 |
| neutrophil mediated immunity | GO:0002446 | 6.46E-13 | 1.10E-09 | MGAM, ARG1, FPR1, PLAUR, PPBP, OLFM4, **MMP8**, ABCA13, ORM2, MMP9, IL6, SELL, TARM1, CXCR2, PRTN3, PGLYRP1, S100A9, **S100A8**, CAMP, CD177, LTF |
| neutrophil degranulation | GO:0043312 | 4.38E-12 | 5.59E-09 | MGAM, ARG1, FPR1, PLAUR, PPBP, OLFM4, **MMP8**, ABCA13, ORM2, MMP9, SELL, TARM1, CXCR2, PRTN3, PGLYRP1, S100A9, **S100A8**, CAMP, CD177, LTF |
| neutrophil activation involved in immune response | GO:0002283 | 5.09E-12 | 5.20E-09 | MGAM, ARG1, FPR1, PLAUR, PPBP, OLFM4, **MMP8**, ABCA13, ORM2, MMP9, SELL, TARM1, CXCR2, PRTN3, PGLYRP1, S100A9, **S100A8**, CAMP, CD177, LTF |
| neutrophil migration | GO:1990266 | 4.55E-11 | 3.87E-08 | CXCR2, CCL4, CCL3, SAA1, PRTN3, CXCL3, S100A9, **S100A8**, CD177 |
| positive regulation of leukocyte chemotaxis | GO:0002690 | 2.29E-09 | 1.67E-06 | CCR1, IL6, SERPINE1, CCL4, CCL3, PPBP, CXCL3, **CXCL2** |
| inflammatory response | GO:0006954 | 2.58E-09 | 1.64E-06 | FPR1, PPBP, CXCL3, **CXCL2**, IL1A, IL6, IL1B, CXCR2, CCL4, CCL3, PROK2, S100A9, **S100A8** |
| neutrophil chemotaxis | GO:0030593 | 2.27E-08 | 1.29E-05 | CXCR2, CCL4, CCL3, SAA1, CXCL3, S100A9, **S100A8** |
| granulocyte chemotaxis | GO:0071621 | 3.37E-08 | 1.72E-05 | CXCR2, CCL4, CCL3, SAA1, CXCL3, S100A9, **S100A8** |
| positive regulation of inflammatory response | GO:0050729 | 2.18E-07 | 1.01E-04 | IL6, SERPINE1, CCL4, CCL3, OSM, S100A9, **S100A8** |
| chemokine-mediated signaling pathway | GO:0070098 | 5.47E-07 | 2.33E-04 | CCR1, CCL4, CCL3, PPBP, CXCL3, **CXCL2** |
| extracellular matrix organization | GO:0030198 | 9.00E-07 | 3.53E-04 | COL1A1, ADAMTS4, VCAN, LOX, SERPINE1, TNC, SPP1, TIMP1, **MMP8**, MMP9 |
| defense response to bacterium | GO:0042742 | 1.43E-06 | 5.21E-04 | SELP, IL6, DMBT1, SERPINE1, REG3G, PGLYRP1, S100A9, **S100A8**, CAMP, LTF |
| cellular response to interleukin-1 | GO:0071347 | 2.66E-06 | 9.05E-04 | IL1A, IL1RN, IL1R2, IL1B, CCL4, CCL3, ACOD1, IL1RAP |
| acute-phase response | GO:0006953 | 3.09E-06 | 9.85E-04 | IL1RN, IL6, IL1B, SAA1 |
| positive regulation of defense response | GO:0031349 | 4.49E-06 | 1.35E-03 | SERPINE1, CCL4, CCL3, S100A9, **S100A8**, EREG |
| regulation of inflammatory response | GO:0050727 | 5.65E-06 | 1.60E-03 | SEMA7A, SERPINE1, CCL4, CCL3, SAA1, ACOD1, S100A9, **S100A8** |
| leukocyte aggregation | GO:0070486 | 6.54E-06 | 1.76E-03 | IL1B, S100A9, **S100A8** |
| antimicrobial humoral immune response mediated by antimicrobial peptide | GO:0061844 | 9.36E-06 | 2.39E-03 | REG3G, PGLYRP1, S100A9, CAMP, LTF |
| positive regulation of response to external stimulus | GO:0032103 | 1.40E-05 | 3.40E-03 | SERPINE1, CCL4, CCL3, ACOD1, S100A9, **S100A8** |
| response to molecule of bacterial origin | GO:0002237 | 2.28E-05 | 5.29E-03 | SELP, IL6, ACOD1, PPBP, CXCL3, **CXCL2** |
| regulation of leukocyte chemotaxis | GO:0002688 | 2.62E-05 | 5.81E-03 | IL6, PPBP, CXCL3, **CXCL2** |
| urea cycle | GO:0000050 | 3.03E-05 | 6.45E-03 | ARG2, ARG1, ACOD1 |
| response to lipopolysaccharide | GO:0032496 | 3.37E-05 | 6.88E-03 | SELP, IL6, SERPINE1, ACOD1, PPBP, CXCL3, **CXCL2** |
| positive regulation of cell proliferation | GO:0008284 | 3.65E-05 | 7.16E-03 | IL11, IL6, IL1B, CXCR2, OSM, PROK2, PRTN3, REG3G, TIMP1, EREG, LTF |
| defense response to Gram-positive bacterium | GO:0050830 | 3.77E-05 | 7.12E-03 | IL6, DMBT1, REG3G, PGLYRP1, CAMP |
| positive regulation of leukocyte migration | GO:0002687 | 4.36E-05 | 7.94E-03 | IL6, PPBP, CXCL3, **CXCL2** |
| positive regulation of cellular process | GO:0048522 | 4.89E-05 | 8.60E-03 | IL11, IL6, IL1B, CXCR2, OSM, PROK2, PRTN3, REG3G, TIMP1, S100A9, **S100A8**, EREG |
| negative regulation of hormone secretion | GO:0046888 | 5.21E-05 | 8.86E-03 | IL11, IL1B, OSM |
| defense response to Gram-negative bacterium | GO:0050829 | 5.77E-05 | 9.51E-03 | SELP, IL6, DMBT1, SERPINE1, LTF |

**Table S2.** 5-fold-CV results of the initial selection. The first three rows achieved the highest AUC and the remaining rows achieved comparable AUC with a simpler model. “Threshold” denotes the probability threshold selected. The standard deviation of the five folds is denoted with ±.

| Panel | Classifier | AUC | Specificity | Sensitivity | Threshold |
| --- | --- | --- | --- | --- | --- |
| CXCL1, IL-12, MMP8, VEGF, S100A8 | Gradient Tree Boosting | $0.98 \pm 0.02$ | $88.9 \pm 4.0$ | $93.1 \pm 6.2$ | 0.49869 |
| CXCL1, MMP8 | Gradient Tree Boosting | $0.97 \pm 0.01$ | $86.0 \pm 9.2$ | $88.9 \pm 7.8$ | 0.49869 |
| CXCL1, IL-12, MMP8 | Gradient Tree Boosting | $0.97 \pm 0.01$ | $85.8 \pm 1.1$ | $89.9 \pm 7.1$ | 0.48535 |
| CXCL2 | Logistic Regression | $0.95 \pm 0.02$ | $83.1 \pm 22.8$ | $89.9 \pm 7.4$ | 0.31535 |
| CXCL1 | Logistic Regression | $0.96 \pm 0.01$ | $75.4 \pm 7.6$ | $95.5 \pm 4.2$ | 0.19525 |
| MMP8 | Logistic Regression | $0.95 \pm 0.03$ | $87.4 \pm 6.0$ | $94.1 \pm 3.1$ | 0.19111 |

**Table S3.**
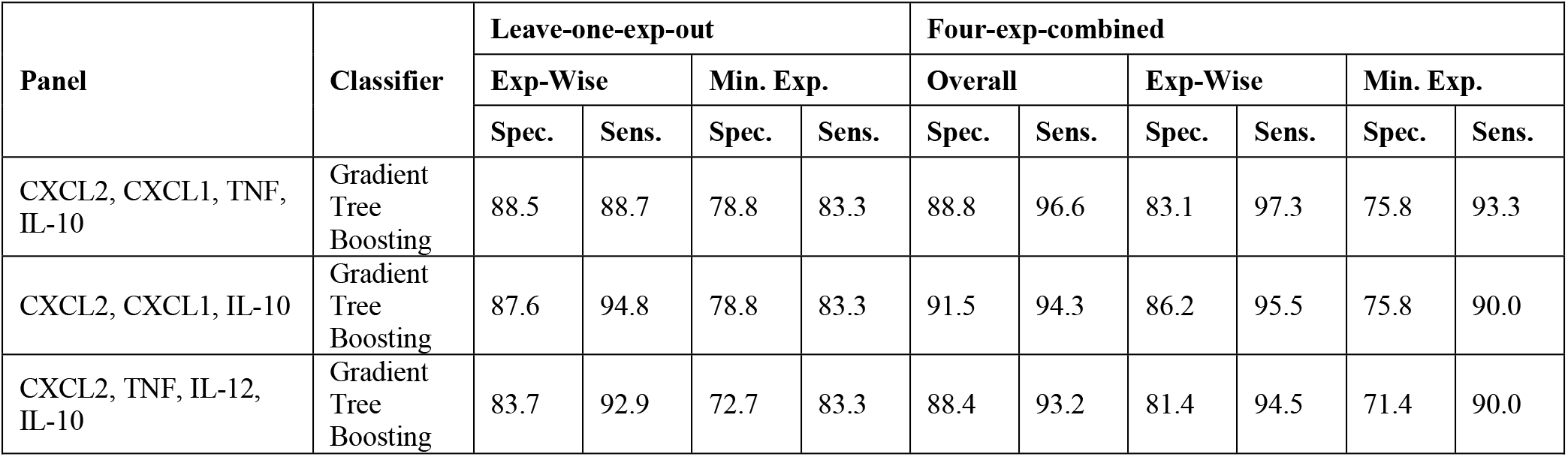
The 100-fold-CV performance of the three classifiers that satisfy the criterion for both leave-one-experiment-out and four-experiments-combined settings. Columns under “Leave-one-exp-out” and “Four-exp-combined” correspond to the performance in leave-one-experiment-out setting and four-experiments-combined setting respectively. “Spec.” denotes specificity and “Sens.” denotes sensitivity. Sensitivity (specificity) under the “Exp-Wise” columns indicate experiment-wise sensitivity (specificity) and sensitivity (specificity) under the “Min. Exp.” columns indicate minimum experiment sensitivity (specificity) and sensitivity (specificity) under the “Overall” columns indicate sensitivity (specificity) (As defined in Materials and methods).

| Panel | Classifier | Leave-one-exp-out |  |  |  | Four-exp-combined |  |  |  |  |  |
| --- | --- | --- | --- | --- | --- | --- | --- | --- | --- | --- | --- |
|  |  | Exp-Wise |  | Min. Exp. |  | Overall |  | Exp-Wise |  | Min. Exp. |  |
|  |  | Spec. | Sens. | Spec. | Sens. | Spec. | Sens. | Spec. | Sens. | Spec. | Sens. |
| CXCL2, CXCL1, TNF, IL-10 | Gradient Tree Boosting | 88.5 | 88.7 | 78.8 | 83.3 | 88.8 | 96.6 | 83.1 | 97.3 | 75.8 | 93.3 |
| CXCL2, CXCL1, IL-10 | Gradient Tree Boosting | 87.6 | 94.8 | 78.8 | 83.3 | 91.5 | 94.3 | 86.2 | 95.5 | 75.8 | 90.0 |
| CXCL2, TNF, IL-12, IL-10 | Gradient Tree Boosting | 83.7 | 92.9 | 72.7 | 83.3 | 88.4 | 93.2 | 81.4 | 94.5 | 71.4 | 90.0 |

**Table S4.** Feature rankings of the ten biomarkers. “Avg.” is short of average and “Diff.” is short of difference. In Materials and Methods, how the average % difference is calculated is described. To calculate the average difference the same method is used but instead of averaging over % difference, it’s averaged over difference. Average AUC denotes the average AUC of the all biomarker panels that contain the biomarker. Average top 5 denotes the average AUC of the five biomarker panels with highest AUC that contain the biomarker and average bottom 5 denotes the average AUC of the five biomarker panels with the least AUC that contain the biomarker.

| Biomarker | Avg. % Diff. | Avg. Diff. | Avg. AUC | Avg. Top 5 | Avg. Bottom 5 |
| --- | --- | --- | --- | --- | --- |
| MMP8 | 11.31 | 0.08 | 0.95 | 0.96 | 0.95 |
| CXCL1 | 5.8 | 0.04 | 0.95 | 0.96 | 0.92 |
| CXCL2 | 4.82 | 0.03 | 0.94 | 0.96 | 0.91 |
| TNF | 3.37 | 0.02 | 0.94 | 0.95 | 0.93 |
| S100A8 | 2.72 | 0.02 | 0.92 | 0.96 | 0.77 |
| CXCL5 | 2.25 | 0.01 | 0.94 | 0.95 | 0.89 |
| IFN-g | 1.14 | 0.01 | 0.93 | 0.95 | 0.83 |
| IL-10 | 0.04 | 0 | 0.92 | 0.96 | 0.67 |
| IL-12 | -0.35 | 0 | 0.92 | 0.96 | 0.65 |
| VEGF | -1.29 | -0.01 | 0.91 | 0.96 | 0.64 |

**Table S5.** The AUC values of the ten biomarkers in the discovery cohort for classifying between SS and nSS mice. 95% confidence intervals are denoted in the parenthesis. AUC values with the confidence intervals were calculated using pROC ^44^.

| Biomarker | AUC |
| --- | --- |
| CXCL5 | 0.89 (0.86-0.93) |
| CXCL2 | 0.96 (0.94-0.99) |
| CXCL1 | 0.97 (0.95-0.98) |
| IFN-g | 0.81 (0.76-0.86) |
| TNF | 0.92 (0.89-0.95) |
| IL-12 | 0.61 (0.55-0.67) |
| IL-10 | 0.66 (0.6-0.71) |
| MMP8 | 0.96 (0.93-0.99) |
| VEGF | 0.58 (0.51-0.65) |
| S100A8 | 0.84 (0.79-0.89) |

**Table S6.** The demographics of the human sera. Patients that are missing the demographic information are omitted while calculating the statistics displayed. For rows next to “Country” and “Sex” the number of patients in each category and its percentage is given. At the final row the median and the IQR of the age is given.

|  |  |  |
| --- | --- | --- |
| Country | South Africa | 47 (49.5%) |
|  | Vietnam | 48 (50.5%) |
| Sex | Female | 34 (35.8%) |
|  | Male | 61 (64.2%) |
| Median Age (IQR) |  | 41 (19.0) |

**Table S7.** (Uploaded as a standalone file.)

The 5-fold-CV AUC values of the 478 classifiers that are searched during the first classifier selection. Highlighted in yellow are the six selected classifiers whose sensitivity and specificity values are reported in Table S2. The standard deviation of the five folds is denoted with ±.

**Table S8.** (Uploaded as a standalone file.)

The 100-fold-CV performance of the 478 classifiers that are searched during the second classifier selection. Highlighted in yellow are the three classifiers that satisfied the criterion. Columns under “Leave-one-exp-out” and “Four-exp-combined” correspond to the performance in the leave-one-experiment-out setting and four-experiments-combined setting respectively. In the leave-one-experiment-out setting, a different experiment is used as the validation set and the columns D-K denote the specificity and sensitivity values on the four validation sets. Columns L and M denote the average of the specificity and sensitivity values in columns D-K respectively. Columns N and O denote the minimum of the specificity and sensitivity values in columns D-K respectively. In the four-experiments-combined setting, four experiments are combined into a dataset. Columns P-W denote the specificity and sensitivity values calculated using only the samples from a single experiment. Columns X and Y denote the average of the specificity and sensitivity values in columns P-W respectively. Columns Z and AA denote the minimum of the specificity and sensitivity values in columns P-W respectively. Columns AB and AC denote the sensitivity and specificity values computed in the usual way i.e. using all of the samples.

### Supplementary Methods

#### Classification algorithm, hyper-parameter and feature selection details

In the first approach, for further analysis, three classifiers with the highest AUC are selected and three classifiers that use a linear decision boundary and a single biomarker which achieved comparable AUCs are selected (Table S2). Operating points of the selected classifiers are tuned to achieve >90% sensitivity and >70% specificity. Of those, we selected the classifier with the highest sensitivity whose specificity is >70% specificity after subtracting one std.

In the second approach, after selecting the hyper-parameters of the classifiers, for each classifier the operating point that maximizes the experiment-wise sensitivity while achieving at least 70% specificity in each of the experiments is selected. Afterward, we identified the classifiers that achieved both minimum experiment sensitivity ≥90% and specificity ≥70 in the four-experiments combined setting and minimum experiment sensitivity ≥80% and specificity ≥70% in the leave-one-experiment-out setting (Table S3). Among the classifiers identified the one with the highest minimum experiment sensitivity (93.3%) is selected.

#### Feature rankings

In the initial classifier selection, we have already measured the AUC of the 239 biomarker combinations using 5-fold-CV on the training portion of the discovery cohort. To calculate the average percent improvement for a biomarker, first, a subset of the 239 biomarker combinations that includes that biomarker are identified. Then, for each identified biomarker combination, the percent difference between the AUC of that combination and the AUC of the biomarker combination without that biomarker is calculated. Finally, percent differences calculated for each of the identified combinations are averaged to calculate the average percent difference for that biomarker. As the AUC of a biomarker combination, we have used the AUC of the Logistic Regression with L1 regularization and the hyper-parameter with the highest AUC is used.

#### Feature rankings plot

The same methodology is used for the initial classifier selection with the exception that all 1023 combinations of the 10 biomarkers are searched and (0.01, 0.3, 0.5,1.) are searched for the hyper-parameters of Gradient Tree Boosting.

#### Logistic Regression with L1 Regularization

We have used the Scikit-learn ^24^ implementation of Logistic Regression with L1 Regularization whose mathematical description is given below. Let ***x***_*i*_ ∈ *R*^*D*^ denote the feature vector and *y*_*i*_ ∈ {−1,1} as the label of i’th sample respectively where D is the number of biomarkers in the panel and {−1,1} denote nSS and SS respectively. Let ***w*** ∈ *R*^*D*^denote the weights of the features and *b* ∈ *R* the bias term, the optimal parameters are found by optimizing the following problem.

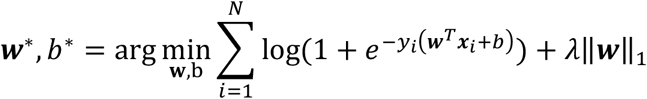

where ‖.‖_1_ denotes the L1 norm.

We have treated *λ* as a hyper-parameter and applied grid search to select it as described in Materials and Methods.

#### Gradient Tree Boosting

Gradient boosting is a way of combining a set of weak learners. Each stage a weak learner is fit to the “pseudo-residuals” resulting from combinations of the weak learners so far. After fitting the weak learner, its prediction function is added to the combined prediction function after multiplied by a shrinkage factor ^45^.

Gradient Tree Boosting uses regression trees as weak learners and uses a slight variation of gradient boosting algorithm where after fitting the regression tree to the “pseudo-residuals” the prediction value for each region is updated again ^45^. The algorithm without the shrinkage is given in Algorithm 10.3 in ^45^.

We have treated maximum depth of the trees, the number of weak learners (referred to as the number of trees), and the shrinkage factor (referred to as the learning rate) as hyper-parameters and selected them through grid search as described in Materials and Methods. The remaining hyper-parameters are used as the default values provided in Scikit-learn ^24^.

### Supplementary Text

#### Selection of the first classifier

Three classifiers achieved the highest area under the receiver operating characteristics curve (AUC) values: Gradient Tree Boosting with CXCL1, IL-12, MMP8, VEGF, and S100A8; Gradient Tree Boosting with CXCL1 and MMP8; and Gradient Tree Boosting with CXCL1, IL-12, and MMP8. The other three classifiers had comparable AUC values and used a single biomarker with a linear decision boundary: Logistic Regression with CXCL2, CXCL1, or MMP8 (Table S2). All six classifiers performed well, with AUCs between 0.95 and 0.98. To refine further, we tuned the operating points to meet the WHO TPP triage test specifications (>90% sensitivity and >70% specificity): Logistic Regression with CXCL1; Logistic Regression with MMP8; and Gradient Tree Boosting with CXCL1, IL-12, MMP8, VEGF, and S100A8. Of those, we selected Logistic Regression with MMP8 for validation, because its sensitivity (94.1%) is higher than that of the Gradient Tree Boosting classifier with five proteins, while its specificity is comparable (87.4%).

#### Selection of the second classifier

In the four-experiment combined setting, 21 out of 478 classifiers achieve minimum experiment sensitivity ≥90 and specificity ≥70. In the leave-one-experiment task, 11 out of the 478 classifiers achieved minimum testing sensitivity ≥80% and specificity ≥70%. None of them achieved higher than 90% minimum testing sensitivity. Three classifiers that satisfy both criterions is listed in (Table S3). We have observed that the number of classifiers achieved sensitivity ≥90 and specificity ≥70 in each of the experiments is less than to the number of classifiers that have experiment wise sensitivity ≥90 and specificity ≥70. We have also observed that the number of classifiers that achieved overall sensitivity ≥90 and specificity ≥70 is higher than the number of classifiers that achieved the same values using experiment-wise averages.

#### Within Experiment Performance of the Selected Classifiers

Logistic regression with MMP8 is evaluated in the testing portion of the discovery cohort in four-experiments combined setting. It achieved sensitivity values within Experiments 1-4, 100%, 100%, 100%, and 89%, respectively and specificity values are 40%, 100%, 70%, and 95% respectively. In the testing portion of the discovery cohort, Gradient Tree Boosting with CXCL1, CXCL2, TNF, and IL-10 is evaluated in four-experiments combined setting. Its sensitivity values within Experiments 1-4 were 87.5%, 100%, 100%, and 100%, respectively, with specificities of 77.8%, 100%, 100%, and 100%, respectively.

## References

1. Organization WH. WHO coronavirus disease (COVID-19) dashboard. 2020. https://covid19.who.int/.

2. WHO. Global Tuberculosis Report Geneva, Switzerland, 2019.

3. Hunter RL, Actor JK, Hwang SA, Karev V, Jagannath C. Pathogenesis of post primary tuberculosis: immunity and hypersensitivity in the development of cavities. Ann Clin Lab Sci 2014; 44(4): 365–87.

4. Levine E. Classification of reinfection pulmonary tuberculosis. The Fundamentals of Pulmonary Tuberculosis and its Complications for Students, Teachers and Practicing Physicians (Hayes E, Ed), Thomas, Springfield 1949: 97–113.

5. Leong FJW-M, Eum S, Via LE, Barry CE, 3rd. Pathology of tuberculosis in the human lung. In: Leong FJ, Dartois V, Dick T, eds. A Color Atlas of Comparative Pathology of Pulmonary Tuberculosis. New York: CRC Press; 2011: 53–81.

6. MacLean E, Broger T, Yerlikaya S, Fernandez-Carballo BL, Pai M, Denkinger CM. A systematic review of biomarkers to detect active tuberculosis. Nat Microbiol 2019; 4(5): 748–58.

7. Ahmad R, Xie L, Pyle M, et al. A rapid triage test for active pulmonary tuberculosis in adult patients with persistent cough. Sci Transl Med 2019; 11(515).

8. Moreira-Teixeira L, Tabone O, Graham CM, et al. Mouse transcriptome reveals potential signatures of protection and pathogenesis in human tuberculosis. Nat Immunol 2020; 21(4): 464–76.

9. WHO. High-priority target product profiles for new tuberculosis diagnostics: report of a consensus meeting, 2014.

10. Churchill GA, Gatti DM, Munger SC, Svenson KL. The Diversity Outbred mouse population. Mamm Genome 2012; 23(9-10): 713–8.

11. Niazi MK, Dhulekar N, Schmidt D, et al. Lung necrosis and neutrophils reflect common pathways of susceptibility to Mycobacterium tuberculosis in genetically diverse, immune-competent mice. Dis Model Mech 2015; 8(9): 1141–53.

12. Harper J, Skerry C, Davis SL, et al. Mouse model of necrotic tuberculosis granulomas develops hypoxic lesions. The Journal of Infectious Diseases 2012; 205(4): 595–602.

13. Smith C, Proulx M, Olive A, et al. Tuberculosis Susceptibility and Vaccine Protection Are Independently Controlled by Host Genotype. mBio 2016; 7(5): e01516.

14. Friedman JH. Greedy function approximation: A gradient boosting machine. Ann Statist 2001; 29(5): 1189–232.

15. Ullman-Cullere MH, Foltz CJ. Body condition scoring: a rapid and accurate method for assessing health status in mice. Lab Anim Sci 1999; 49(3): 319–23.

16. Harrison DE, Astle CM, Niazi MK, Major S, Beamer GL. Genetically diverse mice are novel and valuable models of age-associated susceptibility to Mycobacterium tuberculosis. Immun Ageing 2014; 11(1): 24.

17. Irizarry RA, Hobbs B, Collin F, et al. Exploration, normalization, and summaries of high density oligonucleotide array probe level data. Biostatistics 2003; 4(2): 249–64.

18. Gautier L, Cope L, Bolstad BM, Irizarry RA. affy--analysis of Affymetrix GeneChip data at the probe level. Bioinformatics 2004; 20(3): 307–15.

19. Gentleman RC, Carey VJ, Bates DM, et al. Bioconductor: open software development for computational biology and bioinformatics. Genome Biol 2004; 5(10): R80.

20. Dai M, Wang P, Boyd AD, et al. Evolving gene/transcript definitions significantly alter the interpretation of GeneChip data. Nucleic Acids Research 2005; 33(20): e175–e.

21. Brettschneider J, Collin F, Bolstad BM, Speed TP. Quality Assessment for Short Oligonucleotide Microarray Data. Technometrics 2008; 50(3): 241–64.

22. Benjamini Y, Hochberg Y. Controlling the False Discovery Rate: A Practical and Powerful Approach to Multiple Testing. Journal of the Royal Statistical Society Series B (Methodological) 1995; 57(1): 289–300.

23. Coordinators NR. Database resources of the National Center for Biotechnology Information. Nucleic Acids Research 2012; 41(D1): D8–D20.

24. Pedregosa F, Varoquaux G, Gramfort A, et al. Scikit-learn: Machine Learning in Python. Journal of Machine Learning Research 2011; 12: 2825--30.

25. Kurtz SL, Rossi AP, Beamer GL, Gatti DM, Kramnik I, Elkins KL. The Diversity Outbred Mouse Population Is an Improved Animal Model of Vaccination against Tuberculosis That Reflects Heterogeneity of Protection. mSphere 2020; 5(2).

26. Jasenosky LD, Scriba TJ, Hanekom WA, Goldfeld AE. T cells and adaptive immunity to Mycobacterium tuberculosis in humans. Immunol Rev 2015; 264(1): 74–87.

27. Redford PS, Murray PJ, O’Garra A. The role of IL-10 in immune regulation during M. tuberculosis infection. Mucosal Immunol 2011; 4(3): 261–70.

28. Nouailles G, Dorhoi A, Koch M, et al. CXCL5-secreting pulmonary epithelial cells drive destructive neutrophilic inflammation in tuberculosis. J Clin Invest 2014; 124(3): 1268–82.

29. Jacobs R, Maasdorp E, Malherbe S, et al. Diagnostic Potential of Novel Salivary Host Biomarkers as Candidates for the Immunological Diagnosis of Tuberculosis Disease and Monitoring of Tuberculosis Treatment Response. PLoS One 2016; 11(8): e0160546.

30. Ahmed M, Thirunavukkarasu S, Rosa BA, et al. Immune correlates of tuberculosis disease and risk translate across species. Sci Transl Med 2020; 12(528).

31. Jacobs R, Malherbe S, Loxton AG, et al. Identification of novel host biomarkers in plasma as candidates for the immunodiagnosis of tuberculosis disease and monitoring of tuberculosis treatment response. Oncotarget 2016; 7(36): 57581–92.

32. Wilson D, Badri M, Maartens G. Performance of serum C-reactive protein as a screening test for smear-negative tuberculosis in an ambulatory high HIV prevalence population. PLoS One 2011; 6(1): e15248.

33. Dai Y, Shan W, Yang Q, et al. Biomarkers of iron metabolism facilitate clinical diagnosis in M ycobacterium tuberculosis infection. Thorax 2019; 74(12): 1161–7.

34. Wawrocki S, Seweryn M, Kielnierowski G, Rudnicka W, Wlodarczyk M, Druszczynska M. IL-18/IL-37/IP-10 signalling complex as a potential biomarker for discriminating active and latent TB. PLoS One 2019; 14(12): e0225556.

35. Ruhwald M, Dominguez J, Latorre I, et al. A multicentre evaluation of the accuracy and performance of IP-10 for the diagnosis of infection with M. tuberculosis. Tuberculosis (Edinb) 2011; 91(3): 260–7.

36. Sudbury EL, Otero L, Tebruegge M, et al. Mycobacterium tuberculosis-specific cytokine biomarkers for the diagnosis of childhood TB in a TB-endemic setting. J Clin Tuberc Other Mycobact Dis 2019; 16: 100102.

37. Lin S, Wang Y, Li Y, et al. Diagnostic Accuracy of Interleukin-27 in Bronchoalveolar Lavage Fluids for Pulmonary Tuberculosis. Infect Drug Resist 2019; 12: 3755–63.

38. Manngo PM, Gutschmidt A, Snyders CI, et al. Prospective evaluation of host biomarkers other than interferon gamma in QuantiFERON Plus supernatants as candidates for the diagnosis of tuberculosis in symptomatic individuals. J Infect 2019; 79(3): 228–35.

39. Chegou NN, Sutherland JS, Malherbe S, et al. Diagnostic performance of a seven-marker serum protein biosignature for the diagnosis of active TB disease in African primary healthcare clinic attendees with signs and symptoms suggestive of TB. Thorax 2016; 71(9): 785–94.

40. Albuquerque VVS, Kumar NP, Fukutani KF, et al. Plasma levels of C-reactive protein, matrix metalloproteinase-7 and lipopolysaccharide-binding protein distinguish active pulmonary or extrapulmonary tuberculosis from uninfected controls in children. Cytokine 2019; 123: 154773.

41. Kumar NP, Moideen K, Nancy A, et al. Plasma chemokines are biomarkers of disease severity, higher bacterial burden and delayed sputum culture conversion in pulmonary tuberculosis. Sci Rep 2019; 9(1): 18217.

42. Phalane KG, Kriel M, Loxton AG, et al. Differential expression of host biomarkers in saliva and serum samples from individuals with suspected pulmonary tuberculosis. Mediators Inflamm 2013; 2013: 981984.

43. Jacobs R, Tshehla E, Malherbe S, et al. Host biomarkers detected in saliva show promise as markers for the diagnosis of pulmonary tuberculosis disease and monitoring of the response to tuberculosis treatment. Cytokine 2016; 81: 50–6.

## Supplementary References

44. Robin X, Turck N, Hainard A, et al. pROC: an open-source package for R and S+ to analyze and compare ROC curves. BMC Bioinformatics 2011; 12(1): 77.

45. Hastie T, Tibshirani R, Friedman J. The Elements of Statistical Learning: Data Mining, Inference, and Prediction. New York City, USA: Springer; 2009.

